# Postoperative analgesia and recovery after minimally invasive cardiac surgery

**DOI:** 10.64898/2026.08.27.26361580

**Authors:** Hideaki Note, Takahiro Kajiura, Ai Muramatsu, Yukiko Inagaki, Tetsuro Takahashi, Ko Sato, Kento Nakamura, Takafumi Sadatoshi, Yusuke Sakurai, Masato Tochii, Hirotaka Watanuki, Katsuhiko Matsuyama, Sakura Okamoto

## Abstract

**Introduction:** Postoperative analgesic management after minimally invasive cardiac surgery (MICS) should facilitate early recovery while providing adequate pain control. However, direct evidence comparing postoperative remifentanil-and fentanyl-based analgesic strategies after MICS remains limited. We compared these strategies and explored their associations with postoperative recovery, postoperative nausea and vomiting (PONV), and pain management.

**Methods:** This retrospective single-center observational cohort study included patients who underwent MICS via a right mini-thoracotomy between January 2023 and June 2026. Patients were categorized according to postoperative remifentanil-or fentanyl-based analgesia in the intensive care unit. Outcomes included time to extubation, PONV, postoperative pain assessed using the numerical rating scale (NRS), additional analgesic use, and intensive care unit length of stay. Multivariable logistic regression examined the association between postoperative opioid strategy and PONV, adjusting for age, sex, and smoking history.

**Results:** PONV occurred less frequently in the remifentanil group than in the fentanyl group (20.6% vs 45.0%, P = 0.004), and this association remained significant after adjustment (adjusted odds ratio, 0.23; 95% confidence interval, 0.10–0.56; P = 0.001). Time to extubation was shorter with remifentanil (median, 179 [interquartile range, 134–240.5] vs 247 [190.2–276.5] min; P < 0.001). In contrast, NRS pain scores on postoperative day 0 were higher with remifentanil (3 [1–6] vs 1 [0–2]; P < 0.001), and additional analgesics were used more frequently (80.6% vs 33.3%; P < 0.001). Pain scores on postoperative day 1 did not differ significantly between groups.

**Conclusion:** Postoperative remifentanil-based analgesia after MICS was associated with less PONV and earlier extubation but also with greater early postoperative pain and more frequent additional analgesic use than fentanyl-based analgesia. Appropriate transition to longer-acting analgesics with multimodal analgesia may help preserve the potential benefits of remifentanil while maintaining adequate postoperative pain control.

## Introduction

Current clinical practice guidelines recommend an analgesia-first sedation strategy for mechanically ventilated adult intensive care unit (ICU) patients, making adequate analgesia a fundamental component of contemporary ICU care [1, 2]. These principles are also relevant to patients undergoing cardiac surgery, for whom current Enhanced Recovery After Surgery (ERAS) guidelines recommend multimodal, opioid-sparing analgesia to optimize postoperative recovery [3]. Procedure-specific postoperative analgesic management has therefore become an integral component of contemporary cardiac surgical care.

Minimally invasive cardiac surgery (MICS) was developed to reduce surgical trauma and has been increasingly adopted because it may facilitate faster postoperative recovery than conventional median sternotomy [4]. In this setting, postoperative analgesic management is particularly important because adequate pain control must be achieved while minimizing opioid-related adverse effects that may interfere with early recovery [5]. Although remifentanil and fentanyl have been compared in previous studies of cardiac surgical and general ICU populations [5–7], direct evidence comparing postoperative remifentanil-and fentanyl-based analgesic strategies in the ICU remains limited, particularly in patients undergoing MICS. Consequently, the optimal postoperative opioid-based analgesic strategy after MICS remains uncertain.

We therefore conducted a retrospective cohort study comparing postoperative remifentanil-and fentanyl-based analgesic strategies in patients undergoing MICS. We retrospectively evaluated clinical outcomes related to postoperative recovery, postoperative nausea and vomiting, and pain management to explore their associations with the postoperative opioid-based analgesic strategy.

## Methods

### Study design

This retrospective single-center observational cohort study was conducted at Aichi Medical University Hospital. Patients who underwent minimally invasive cardiac surgery (MICS) between January 2023 and June 2026 were retrospectively identified from the institutional electronic medical records and categorized according to the postoperative opioid administered in the intensive care unit (ICU) (remifentanil or fentanyl). The study protocol was approved by the Institutional Review Board of Aichi Medical University Hospital (approval No. 2024-220) and was conducted in accordance with the principles of the Declaration of Helsinki. The requirement for individual written informed consent was waived because of the retrospective nature of the study. Instead, an opt-out approach was adopted by publicly disclosing the study information and offering patients the opportunity to decline participation.

### Study population

Adult patients who underwent minimally invasive cardiac surgery (MICS) via a right mini-thoracotomy at Aichi Medical University Hospital between January 2023 and June 2026 were eligible for inclusion. Surgical procedures included mitral valve repair or replacement, aortic valve replacement, tricuspid valve surgery, atrial septal defect closure, cardiac myxoma resection, and other cardiac procedures performed through the same approach. Patients who received postoperative analgesia using either a fentanyl-based or remifentanil-based strategy in the intensive care unit (ICU) were included.

Patients were excluded if conversion to median sternotomy or any reoperation was required, or if major postoperative complications substantially altered the expected postoperative clinical course and precluded reliable assessment of the study outcomes. Examples included severe respiratory or hemodynamic complications requiring prolonged ventilatory or circulatory support. If a patient underwent more than one eligible MICS procedure during the study period, only the first operation was included in the analysis. The patient selection process is shown in Figure 1.

**Fig 1.**
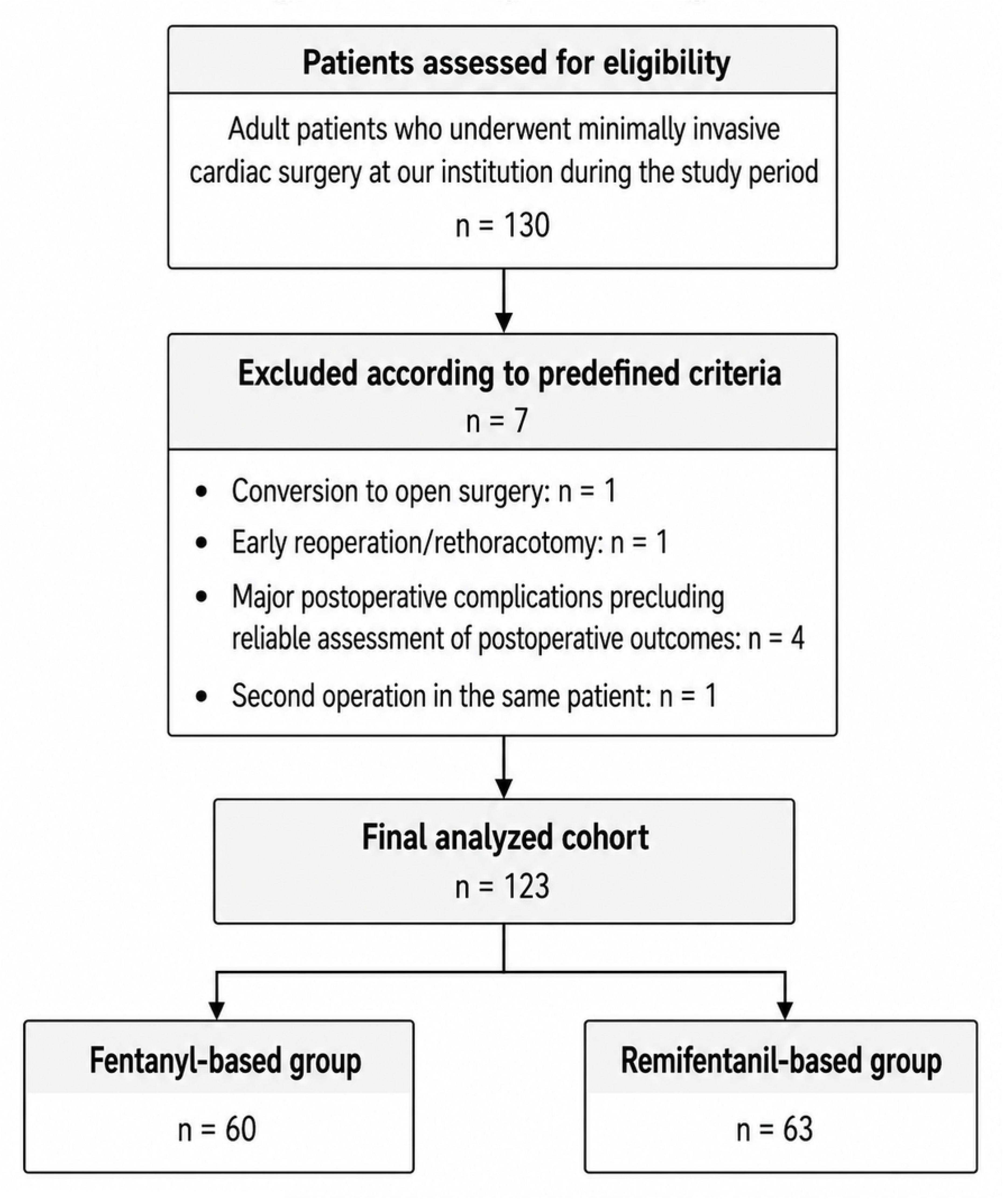
Study flow diagram.

Among 130 adult patients who underwent minimally invasive cardiac surgery during the study period, 7 were excluded because of conversion to open surgery, early reoperation/rethoracotomy, major postoperative complications precluding reliable assessment of postoperative outcomes, or duplicate surgery in the same patient. The final analyzed cohort consisted of 123 patients, including 60 in the fentanyl-based group and 63 in the remifentanil-based group.

### Perioperative management

All patients underwent MICS via a right mini-thoracotomy under general anesthesia and were admitted to the ICU after surgery. General anesthesia was induced primarily with propofol, with midazolam used in selected patients at the discretion of the attending anesthesiologist. Remifentanil, fentanyl, and rocuronium were administered intraoperatively in all patients. Anesthesia was maintained with sevoflurane before cardiopulmonary bypass (CPB), switched to propofol-based total intravenous anesthesia during CPB, and returned to sevoflurane as appropriate after weaning from CPB. Postoperative analgesia and sedation in the ICU were managed by the treating ICU physicians according to each patient’s clinical condition. Patients received either a remifentanil-based or fentanyl-based postoperative analgesic strategy. Infusion rates were approximately 0.07–0.10 μg/kg/min for remifentanil and 0.4–0.5 μg/kg/h for fentanyl, with subsequent dose adjustments based on clinical requirements. Propofol and dexmedetomidine were used as needed for postoperative sedation. In preparation for extubation, remifentanil and propofol were discontinued, whereas fentanyl and dexmedetomidine could be continued, with dose reduction as clinically appropriate. Acetaminophen was routinely administered to all patients. Additional analgesics, including nonsteroidal anti-inflammatory drugs (NSAIDs), tramadol, and mirogabalin, were administered as clinically indicated. Regional analgesic techniques were not performed. Tracheal extubation was performed when clinically appropriate, as determined by the treating ICU physicians based on the patient’s overall clinical condition.

### Outcome measures

Clinical outcomes related to postoperative recovery, postoperative nausea and vomiting (PONV), and pain management were retrospectively evaluated to explore their associations with the postoperative opioid-based analgesic strategy. Data were extracted from the electronic medical records by the study investigators, including anesthesia records, ICU records, nursing records, and medication administration records.

The electronic medical records were accessed for research purposes from January 6, 2025 to July 14, 2026. During data collection, the investigators had access to information that could identify individual patients. The analytic dataset was subsequently de-identified, and only de-identified data were used thereafter.

Time to extubation was defined as the interval, in minutes, from ICU admission to tracheal extubation. Patients who did not undergo extubation because of reoperation or major postoperative complications were excluded as described above. ICU length of stay was calculated as the number of days from ICU admission to ICU discharge.

PONV was defined as the occurrence of either nausea or vomiting documented in the electronic medical records, including nursing records, from extubation through postoperative day 1 (POD1). Nausea and vomiting were analyzed together as a composite PONV outcome.

Postoperative pain was assessed using the numerical rating scale (NRS), ranging from 0 (no pain) to 10 (worst pain imaginable), on POD0 and POD1. When multiple NRS measurements were available within the same postoperative day, the highest recorded value was used for analysis. NRS assessments were not differentiated between pain at rest and pain during movement. Additional analgesic use was defined as the administration of NSAIDs, tramadol, or mirogabalin and was evaluated as a binary outcome (use or no use). The use of each analgesic was also evaluated separately. Oral intake on POD1 and the occurrence of delirium were recorded as binary postoperative outcomes.

### Statistical analysis

Continuous variables are presented as medians with interquartile ranges (IQRs), and categorical variables as numbers and percentages. For baseline patient and perioperative characteristics, continuous variables were compared between the fentanyl-based and remifentanil-based groups using the Mann–Whitney U test. Binary categorical variables were compared using Fisher’s exact test, and the distribution of surgical procedure categories was compared using Pearson’s chi-square test.

Postoperative outcomes were analyzed using available cases for each outcome. Continuous or ordinal variables were compared using the Mann–Whitney U test. Categorical variables were compared using Pearson’s chi-square test; Fisher’s exact test was used when any expected cell count was less than 5.

To further examine the association between opioid regimen and postoperative nausea and vomiting (PONV), univariable and multivariable logistic regression analyses were performed. The multivariable model included opioid group, age, sex, and smoking history. Age was modeled per 10-year increase. Odds ratios (ORs) with 95% confidence intervals (CIs) were calculated. Univariable analyses were performed using available cases, whereas the multivariable analysis was restricted to complete cases because smoking history was missing for one patient.

All statistical tests were two-sided, and a P value <0.05 was considered statistically significant. Statistical analyses were performed using R version 4.5.1 (R Foundation for Statistical Computing, Vienna, Austria).

## Results

### Patient and perioperative characteristics

A total of 123 patients were included in the analysis, comprising 60 patients in the fentanyl-based group and 63 in the remifentanil-based group. Patient and perioperative characteristics are summarized in Table 1. Patients in the remifentanil-based group were younger than those in the fentanyl-based group (median age, 63 [IQR, 52.5–72.5] vs 73 [56.8–80] years; P = 0.004). Aortic cross-clamp time was also shorter in the remifentanil-based group (95 [80–109] vs 104 [95–124] min; P = 0.010). Other baseline characteristics, comorbidities, surgical procedures, and intraoperative variables did not differ significantly between the groups.

**Table 1.**
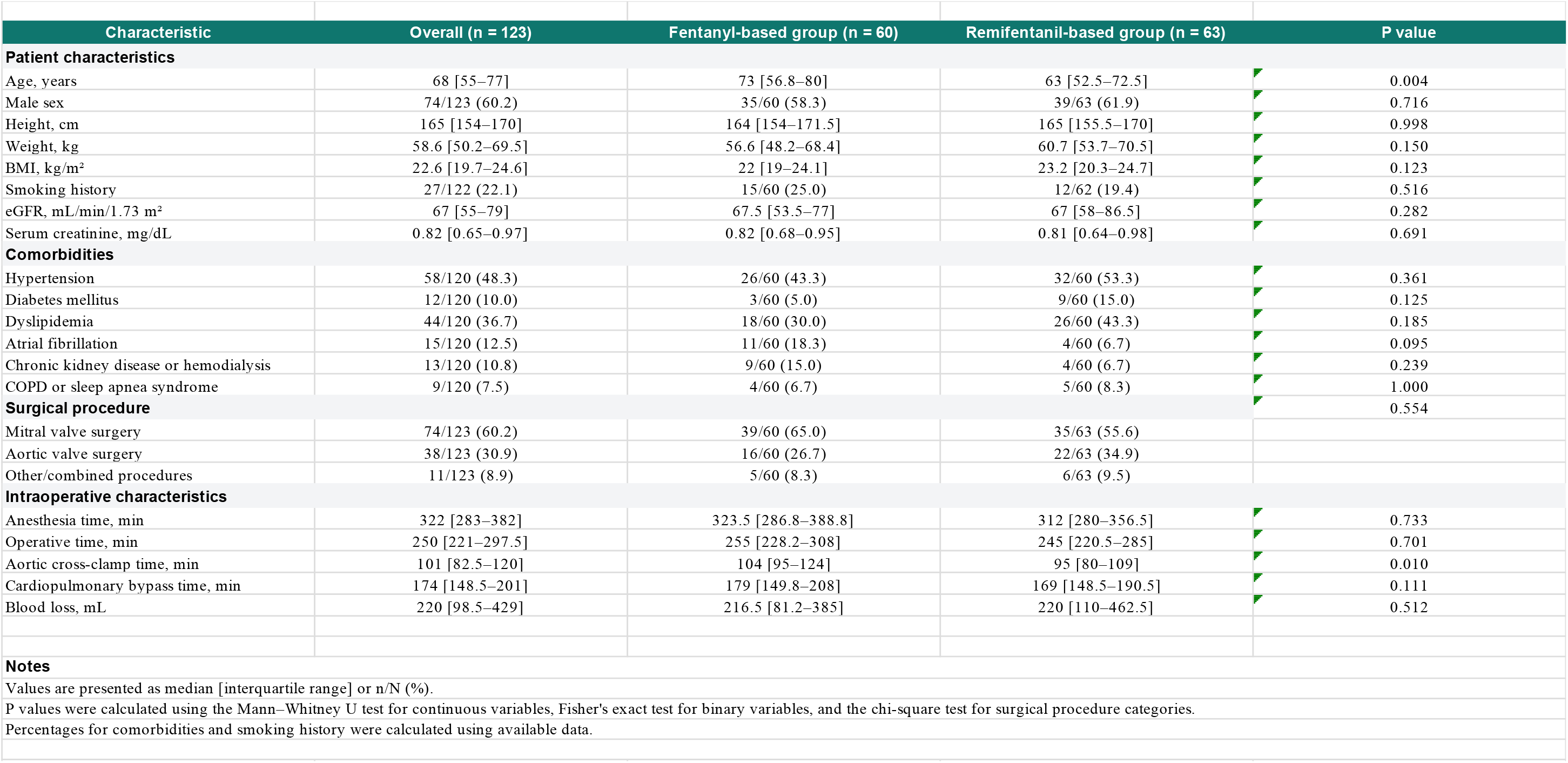
Patient and perioperative characteristics.

### Postoperative outcomes

Postoperative outcomes are shown in Table 2. PONV occurred less frequently in the remifentanil-based group than in the fentanyl-based group (20.6% vs 45.0%; P = 0.004). In contrast, the NRS pain score on postoperative day 0 was higher in the remifentanil-based group (median, 3 [IQR, 1–6] vs 1 [0–2]; P < 0.001), whereas no significant difference was observed on postoperative day 1 (3 [0.5–5.5] vs 3 [0–5]; P = 0.218).

**Table 2.** Postoperative outcomes in the analyzed cohort.

| Outcome | Fentanyl-based group (n=60) | Remifentanyl-based group (n=63) | p value | Statistical test | Available n (F/R) |
| --- | --- | --- | --- | --- | --- |
| Postoperative nausea and vomiting, n (%) | 27/60 (45.0%) | 13/63 (20.6%) | 0.004 | Chi-square test | 60/63 |
| NRS pain score on postoperative day 0, median [IQR] | 1 [0–2] | 3 [1–6] | <0.001 | Mann–Whitney U test | 60/63 |
| NRS pain score on postoperative day 1, median [IQR] | 3 [0–5] | 3 [0.5–5.5] | 0.218 | Mann–Whitney U test | 60/63 |
| Any additional analgesic use, n (%) | 20/60 (33.3%) | 50/62 (80.6%) | <0.001 | Chi-square test | 60/62 |
| NSAID use, n (%) | 15/60 (25.0%) | 46/63 (73.0%) | <0.001 | Chi-square test | 60/63 |
| Tramadol use, n (%) | 12/60 (20.0%) | 27/62 (43.5%) | 0.005 | Chi-square test | 60/62 |
| Mirogabalin use, n (%) | 0/60 (0.0%) | 39/63 (61.9%) | <0.001 | Chi-square test | 60/63 |
| Oral intake on postoperative day 1, n (%) | 45/60 (75.0%) | 56/63 (88.9%) | 0.045 | Chi-square test | 60/63 |
| Time from ICU admission to extubation, min, median [IQR] | 247 [190.2–276.5] | 179 [134–240.5] | <0.001 | Mann–Whitney U test | 60/60 |
| ICU stay, days, median [IQR] | 3 [3–3] | 3 [3–3] | 0.769 | Mann–Whitney U test | 60/63 |
| Delirium, n (%) | 1/60 (1.7%) | 1/63 (1.6%) | 1.000 | Fisher exact test | 60/63 |
| **Notes** |  |  |  |  |  |
| Values are presented as n/N (%) or median [interquartile range]. |  |  |  |  |  |
| Categorical variables were compared using Pearson’s chi-square test unless expected cell counts were <5, in which case Fisher’s exact test was used. |  |  |  |  |  |
| Continuous or ordinal variables were compared using the Mann–Whitney U test. |  |  |  |  |  |
| F/R: available observations in the fentanyl-based/remifentanyl-based groups. |  |  |  |  |  |

Additional analgesic use was more frequent in the remifentanil-based group (80.6% vs 33.3%; P < 0.001). The use of NSAIDs (73.0% vs 25.0%; P < 0.001), tramadol (43.5% vs 20.0%; P = 0.005), and mirogabalin (61.9% vs 0%; P < 0.001) was also more frequent in the remifentanil-based group.

Oral intake on postoperative day 1 was more frequent in the remifentanil-based group (88.9% vs 75.0%; P = 0.045). The time from ICU admission to extubation was shorter in the remifentanil-based group (median, 179 [IQR, 134–240.5] vs 247 [190.2–276.5] min; P < 0.001). No patient included in the analysis required reintubation after extubation. ICU length of stay and the incidence of delirium did not differ significantly between the groups.

### Factors associated with PONV

The association between opioid regimen and PONV was further examined using logistic regression analysis (Table 3). In univariable analysis, the remifentanil-based regimen was associated with lower odds of PONV compared with the fentanyl-based regimen (OR, 0.32; 95% CI, 0.14–0.70; P = 0.005).

**Table 3.** Factors associated with postoperative nausea and vomiting.

| Variable | Univariable OR (95% CI) | P value | Adjusted OR (95% CI) | P value |
| --- | --- | --- | --- | --- |
| Remifentanyl-based group (vs fentanyl-based group) | 0.32 (0.14–0.70) | 0.005 | 0.23 (0.10–0.56) | 0.001 |
| Age, per 10-year increase | 0.93 (0.71–1.23) | 0.623 | 0.77 (0.56–1.04) | 0.088 |
| Female sex (vs male) | 1.86 (0.87–4.01) | 0.112 | 1.97 (0.82–4.73) | 0.130 |
| Smoking history (yes vs no) | 0.69 (0.26–1.80) | 0.447 | 0.69 (0.24–1.98) | 0.493 |
| *Values are odds ratios with 95% confidence intervals.* |  |  |  |  |
| *Multivariable logistic regression included group, age, sex, and smoking history.* |  |  |  |  |
| *Univariable analyses used available cases; multivariable analysis was performed in complete cases (n = 122) because smoking history was missing in one patient.* |  |  |  |  |
| *Outcome: postoperative nausea and vomiting.* |  |  |  |  |

In the multivariable logistic regression model including opioid group, age, sex, and smoking history, the remifentanil-based regimen remained significantly associated with lower odds of PONV. After adjustment for age, sex, and smoking history, the adjusted OR for PONV in the remifentanil-based group was 0.23 (95% CI, 0.10–0.56; P = 0.001). Age, sex, and smoking history were not significantly associated with PONV in the multivariable model. The multivariable analysis included 122 complete cases because smoking history was missing for one patient.

## Discussion

### Summary of the main findings

In this retrospective study of patients undergoing MICS, postoperative remifentanil-based analgesia was associated with a lower incidence of postoperative nausea and vomiting and earlier extubation compared with fentanyl-based analgesia. Patients receiving remifentanil experienced greater early postoperative pain and required additional analgesics more frequently.

#### PONV

The lower incidence of postoperative nausea and vomiting (PONV) in the remifentanil group remained significant after adjustment for patient characteristics, suggesting that this association was not solely explained by baseline differences between the groups. Current PONV guidelines recognize perioperative opioid exposure as one of the major modifiable risk factors for PONV and recommend opioid-sparing analgesic strategies whenever feasible [8]. Consistent with our findings, Rama-Maceiras et al. reported a lower incidence of PONV in patients receiving propofol–remifentanil anesthesia than in those receiving propofol–fentanyl anesthesia during plastic surgery [9]. In contrast, Choi et al. found that intraoperative remifentanil infusion was independently associated with an increased risk of PONV in patients receiving fentanyl-based intravenous patient-controlled analgesia [10]. This apparent discrepancy may be explained by differences in postoperative opioid exposure. In the study by Choi et al., remifentanil was used only intraoperatively and was followed by postoperative fentanyl-based analgesia, whereas our study compared two postoperative ICU analgesic strategies. Therefore, patients in the remifentanil group were likely exposed to less residual opioid during the early postoperative period. Taken together, these findings suggest that differences in residual opioid exposure and postoperative analgesic strategy, rather than remifentanil itself, may have contributed to the lower incidence of PONV observed in our study.

### Early extubation

Earlier extubation was observed in the remifentanil group. This finding is biologically plausible because remifentanil is rapidly metabolized by nonspecific plasma and tissue esterases, resulting in an extremely short context-sensitive half-life with minimal accumulation even after prolonged infusion. Consequently, its analgesic and respiratory depressant effects resolve rapidly after discontinuation, whereas fentanyl may continue to exert residual respiratory depressant effects. These pharmacokinetic characteristics have been associated with faster emergence and shorter duration of mechanical ventilation in systematic reviews of ICU sedation and general anesthesia [11, 12].

Previous studies evaluating the effect of remifentanil on extubation have reported inconsistent findings. Two recent network meta-analyses found no significant reduction in the duration of mechanical ventilation with remifentanil compared with fentanyl or other opioids after cardiac surgery or in mechanically ventilated ICU patients [6, 13]. In contrast, Muellejans et al. demonstrated significantly earlier extubation with a remifentanil/propofol-based sedation regimen than with conventional midazolam/fentanyl sedation after cardiac surgery [14]. However, another randomized trial comparing remifentanil-and fentanyl-based ICU analgesia found no significant difference in extubation time [7]. Collectively, these findings suggest that the impact of remifentanil on extubation remains uncertain and is likely influenced by differences in patient populations, perioperative management, and postoperative analgesic protocols. One possible explanation for our findings is that the relatively homogeneous population undergoing MICS allowed differences in postoperative analgesic strategies to become more apparent. Early extubation is a fundamental component of enhanced recovery pathways after MICS, and optimization of postoperative analgesic strategy may therefore exert a greater influence on extubation timing than in conventional cardiac surgery, where recovery is affected by a broader range of procedure-related factors.

However, our findings should be interpreted with caution. Patients in the fentanyl group were older than those in the remifentanil group, and age may have contributed to the observed difference in extubation time. Furthermore, multivariable adjustment was not performed for this outcome. Therefore, the association between postoperative remifentanil-based analgesia and earlier extubation should be considered exploratory rather than definitive, and prospective studies are warranted to determine whether remifentanil independently facilitates earlier extubation after MICS.

### Postoperative pain

Patients receiving postoperative remifentanil-based analgesia experienced greater early postoperative pain and required additional analgesics more frequently than those receiving fentanyl. Direct comparative evidence regarding postoperative pain following postoperative remifentanil-versus fentanyl-based ICU analgesia after cardiac surgery remains limited. To our knowledge, no previous study has specifically compared these two postoperative ICU analgesic strategies with postoperative pain as the primary outcome.

One plausible explanation is the rapid loss of analgesic effect after discontinuation of remifentanil. Because remifentanil has an ultra-short context-sensitive half-life, inadequate transition to longer-acting analgesics before discontinuation may create an immediate postoperative analgesic gap, resulting in greater early postoperative pain and increased need for additional analgesia [15]. This interpretation is supported by De Hoogd et al., who reported that patients receiving intraoperative remifentanil required significantly more postoperative morphine during the first 48 hours after cardiac surgery than those receiving fentanyl, although remifentanil was administered only intraoperatively in that study [16].

Another possible explanation is the development of acute opioid tolerance or opioid-induced hyperalgesia associated with remifentanil exposure. Experimental and clinical studies have suggested that these phenomena may occur, particularly after high-dose administration or abrupt discontinuation of remifentanil [15]. In addition, a recent systematic review and meta-analysis demonstrated that intravenous magnesium attenuated remifentanil-induced postoperative hyperalgesia, supporting the clinical relevance of this mechanism [17]. However, our study was not designed to distinguish analgesic gap from opioid-induced hyperalgesia or acute opioid tolerance, and the contribution of these mechanisms to the observed increase in postoperative pain remains uncertain.

In contrast, Subramaniam et al. found no significant differences in postoperative pain scores or opioid consumption between intraoperative remifentanil and fentanyl during cardiac surgery [18]. Their findings suggest that postoperative pain may be influenced not only by the pharmacological properties of remifentanil but also by perioperative analgesic management. Differences in transition analgesia, multimodal analgesia, and postoperative pain management protocols may therefore modify the clinical impact of remifentanil on postoperative pain.

However, these findings should be interpreted with caution. Multivariable adjustment was not performed for postoperative pain, and differences in concomitant analgesics and postoperative management between the groups may have influenced pain intensity and rescue analgesic requirements. Therefore, the association between postoperative remifentanil-based analgesia and greater early postoperative pain should be considered exploratory rather than definitive.

### Clinical implications

The present findings suggest that postoperative remifentanil-based analgesia may facilitate early postoperative recovery but may also increase early postoperative pain, highlighting an important trade-off in postoperative analgesic management after MICS. This trade-off may be mitigated by appropriate transition to longer-acting analgesics together with multimodal analgesia. Accordingly, postoperative opioid selection should be considered as one component of an integrated enhanced recovery strategy after MICS, tailored to individual patient characteristics and perioperative management.

### Limitations

This study has several limitations. First, its retrospective single-center design limits causal inference, and baseline differences between the study groups may have influenced the observed outcomes. Second, extubation timing, postoperative analgesic management, and additional analgesic administration were determined by the treating clinicians rather than by standardized protocols, introducing potential practice variation. Third, changes in perioperative practice over the study period may have influenced the observed outcomes. Finally, because of the observational design, it was not possible to distinguish the effects of the opioid itself from those of the overall postoperative analgesic strategy. Prospective multicenter studies using standardized extubation and analgesic protocols are warranted to confirm these findings and to establish the optimal postoperative analgesic strategy after MICS.

## Conclusion

Postoperative remifentanil-based analgesia after MICS was associated with a lower incidence of postoperative nausea and vomiting and earlier extubation but also with greater early postoperative pain and more frequent use of additional analgesics compared with fentanyl-based analgesia. To maximize the benefits of postoperative remifentanil-based analgesia after MICS, appropriate transition to longer-acting analgesics together with multimodal analgesia should be incorporated into postoperative analgesic management.

## Data Availability

The data underlying this study contain potentially identifying and sensitive patient information and therefore cannot be made publicly available. De-identified data may be made available to qualified researchers upon reasonable request and in accordance with applicable ethical and institutional requirements. Requests for data access should be directed to the Ethics Committee Office of Aichi Medical University.

## Acknowledgments

None.

## Supporting information

S1 Table. Full results of the univariable and multivariable logistic regression analyses for postoperative nausea and vomiting.

## Notes

### Competing Interest Statement

The authors have declared no competing interest.

### Author Declarations

The study protocol was approved by the Institutional Review Board of Aichi Medical University Hospital (approval No. 2024-220) and was conducted in accordance with the principles of the Declaration of Helsinki. The requirement for individual written informed consent was waived because of the retrospective nature of the study. Instead, an opt-out approach was adopted by publicly disclosing the study information and offering patients the opportunity to decline participation.

